# Poisonings and Lifetime Use of Nitrous Oxide in the United States from 2021 to 2023

**DOI:** 10.64898/2025.12.23.25342906

**Authors:** Orrin D. Ware

## Abstract

**Introduction:** Harms associated with recreational nitrous oxide use have been documented. Studies on the prevalence of past-year inhalant use disorder among individuals who have ever used nitrous oxide are lacking. This descriptive study, conducted in the United States from 2021 to 2023, examined the prevalence of ever using nitrous oxide, determined the proportion of people meeting the past-year criteria for an inhalant use disorder, and reported the yearly number of nitrous oxide poisonings.

**Methods:** This study used a descriptive epidemiological approach to identify and characterize the lifetime use of nitrous oxide and the total number of nitrous oxide poisonings as reported in the United States from 2021 to 2023.

**Results:** From 2021 to 2023, one in twenty people in the U.S. reported ever using nitrous oxide recreationally. The prevalence of inhalant use disorder in the past year among those who ever used nitrous oxide was 1.1%. However, this diagnosis could be due to the use of other inhalants such as poppers. Among individuals who only used nitrous oxide in their lifetime to the exclusion of all other inhalants, and who reported using inhalants in the past year, approximately 10% met the criteria for inhalant use disorder due to their nitrous oxide use. During this period, there were 937 total poisoning cases involving nitrous oxide, with 712 cases involving nitrous oxide as the only reported substance.

**Conclusions:** More population-level studies are needed to examine the national harms of problematic nitrous oxide use.

## INTRODUCTION

The dissociative anesthetic with analgesic properties, nitrous oxide (N_2_O), also known as whippets or whippits, and sometimes called "hippy crack” or "nangs," is misused recreationally.^1-6^ According to the 2019 Global Drug Survey, it ranks among the top fifteen substances ever reported as used in respondents’ lifetime, with 23.5% of the sample endorsing its use and 12% reporting use in the past 12 months.^7^ Besides being an anesthetic, nitrous oxide has commercial purposes, such as its use in the food industry in whipped cream cans, which some individuals misuse to become intoxicated.^2, 4, 8, 9^ Recreational use of nitrous oxide has also been reported at social events like clubs, festivals, and parties,^10-13^ and it has a history of misuse by some healthcare providers with access to it.^5^ With a rapid onset, the psychoactive effects of nitrous oxide are relatively brief, usually lasting a few minutes or less.^1, 2, 6^ With the short duration of its effects, some individuals choose to use the substance multiple times.^1, 3, 6, 8, 14^

Harms and risks associated with problematic or long-term use of nitrous oxide include B12 deficiency,^10, 15, 16^ emphysema,^9^ neurological impacts such as neuropathy,^3^ psychiatric symptoms such as anxiety or depression,^5, 17^ and driving under the influence.^2, 6, 18^ The problematic use of nitrous oxide could lead to individuals developing a substance use disorder, specifically a diagnosis of an “other substance use disorder”.^2-4, 9, 19^ It is also important to note that nitrous oxide is sometimes classified as an inhalant, contributing to its use disorder being classified as an inhalant use disorder in some epidemiological datasets. Two such examples in the United States are the National Survey on Drug Use and Health (NSDUH), a nationally representative health survey, and the Treatment Episode Data Set, a real-world dataset of substance use disorder treatment admissions and discharges.^20, 21^ This is an essential consideration as withdrawal is not a symptom that is diagnostically present for inhalant or hallucinogen drug classes based on the Diagnostic and Statistical Manual of Mental Disorders: Fifth Edition, Text Revision.^19^ As a dissociative anesthetic that is inhaled, the withdrawal symptom associated with substance use disorders, such as alcohol use disorder or opioid use disorder, are not commonly associated with nitrous oxide use.^2, 3, 22^

Globally, males, young adults, and adolescents tend to misuse nitrous oxide the most.^9, 17^ Globally, harms have also been identified related to nitrous oxide use, including several case reports, with its use being a precipitating factor in poisonings, emergency department visits, and deaths.^2, 17, 23^ The reported prevalence of ever using nitrous oxide in the United States was approximately 2% in 2002 and 2003, 4.6% in 2015, and 4.1% in 2018.^13, 24, 25^ However, there is a paucity of studies that focus on population-level recreational nitrous oxide use.^1, 18^ Further, population-level studies examining meeting the criteria for an inhalant use disorder among persons who have ever used nitrous oxide are also lacking.^2^ Therefore, the purpose of this descriptive study, conducted in the U.S. from 2021 to 2023, was threefold: [1] to determine the prevalence of lifetime nitrous oxide use, [2] to assess how many individuals who ever used nitrous oxide met the criteria for an inhalant use disorder in the past twelve months, and [3] to examine the annual number of nitrous oxide poisonings.

## METHODS

### Data Source

Cross-sectional data from 2021 to 2023 from the Annual Report of the National Poison Data System® (NPDS) from America’s Poison Centers®^26-28^ and NSDUH^20^ were used for this study. The NSDUH 2021 to 2023 is a de-identified concatenated cross-sectional file that provides annual data from a national health survey among a nationally representative sample in the U.S. The NSDUH is a household survey in which respondents are asked about health-related topics, including substance use. The Annual Report of the NPDS provides annual data from calls and reported exposures to poison centers in the U.S. The Annual Report of the NPDS contains total counts of specific substances being reported for a specific year such as nitrous oxide.

### Sample Selection

There are 173,808 unweighted cases in the NSDUH 2021 to 2023 concatenated file. Cases were selected for inclusion in this study if they reported ever inhaling nitrous oxide or whippits based on a survey item that directly asked respondents. Cases were also selected if they reported ever using “Nitrous oxide, "whippits," dentist gas”, “Whipped-cream can”, or “Hippie crack” when asked to report other substances they have used. This resulted in an unweighted sample of 7,372 cases across the analytic period from 2021 to 2023. Nitrous oxide poisoning data extracted from the Annual Report of the NPDS include: number of case mentions (all cases in which nitrous oxide was involved), number of single case mentions (cases in which nitrous oxide was the only contributing substance, and intentional cases (which includes the intentional use such as misuse, self-harm, recreational use, and medical error).

### Analysis

This study employed a descriptive epidemiological approach to identify and characterize this public health phenomenon instead of significance testing.^29, 30^ SPSS Version 31^31^ was used, and study procedures were considered not human subjects research based on the University of North Carolina at Chapel Hill Institutional Review Board. Descriptive statistics such as counts and percentages were used. After selecting the sample from the NSDUH, one-year sample weights were applied. Factors examined in the NSDUH data include: year of data collection, age group, race and ethnicity, sex, whether the respondent ever used other specific inhalants such as poppers, correction fluid, gasoline, or spray paints, time since last used inhalants, and whether the respondent met the criteria for a past year inhalant use disorder. Individuals who met the criteria for a past year inhalant use disorder were then identified as whether their inhalant use disorder was mild, moderate or severe. To identify cases in which inhalant use disorder was only attributable to nitrous oxide instead of other inhalants, a sub-sample of cases was selected based on the following criteria: [a] lifetime use of nitrous oxide was reported, [b] past 12 month inhalant use was reported, and [c] the respondent never used other inhalants. Data from the Annual Report of the NPDS were presented as counts in this current study.

## RESULTS

### Lifetime Use of Nitrous Oxide: 2021 to 2023

After applying the one-year sample weights, the selected sample of 7,372 cases from the NSDUH data represented 39,376,936 persons. This accounted for 4.7% of the U.S. population during the entire analytic period. Therefore, among the combined sample of the U.S. population from 2021 to 2023, approximately 4.7% ever used nitrous oxide. Annually, 4.4% (12,299,426 individuals), 4.9% (13,733,955 individuals), and 4.7% (13,343,555 individuals) of the population reported ever using nitrous oxide from 2021, 2022, and 2023, respectively. Table 1 presents demographic characteristics, inhalant use characteristics, and past-year inhalant use disorder characteristics across the analytic sample. As seen in Table 1, the sample was primarily Non-Hispanic White at 83.1% (n = 32,725,647) and male at 64.6% (n = 25,453,475). The largest age group were 35-49 year olds at 37.9%. Nearly 30% of the sample, ever used amyl nitrates or poppers (n = 11,557,241). The majority of the sample last used inhalants more than 12 months ago at 92.2% (n = 36,295,925), 5.3% (n = 2,097,956) used inhalants with the past 12 months but more than 30 days ago, and 2.1% (n = 828,168) used inhalants in the past 30 days.

**Table 1.**
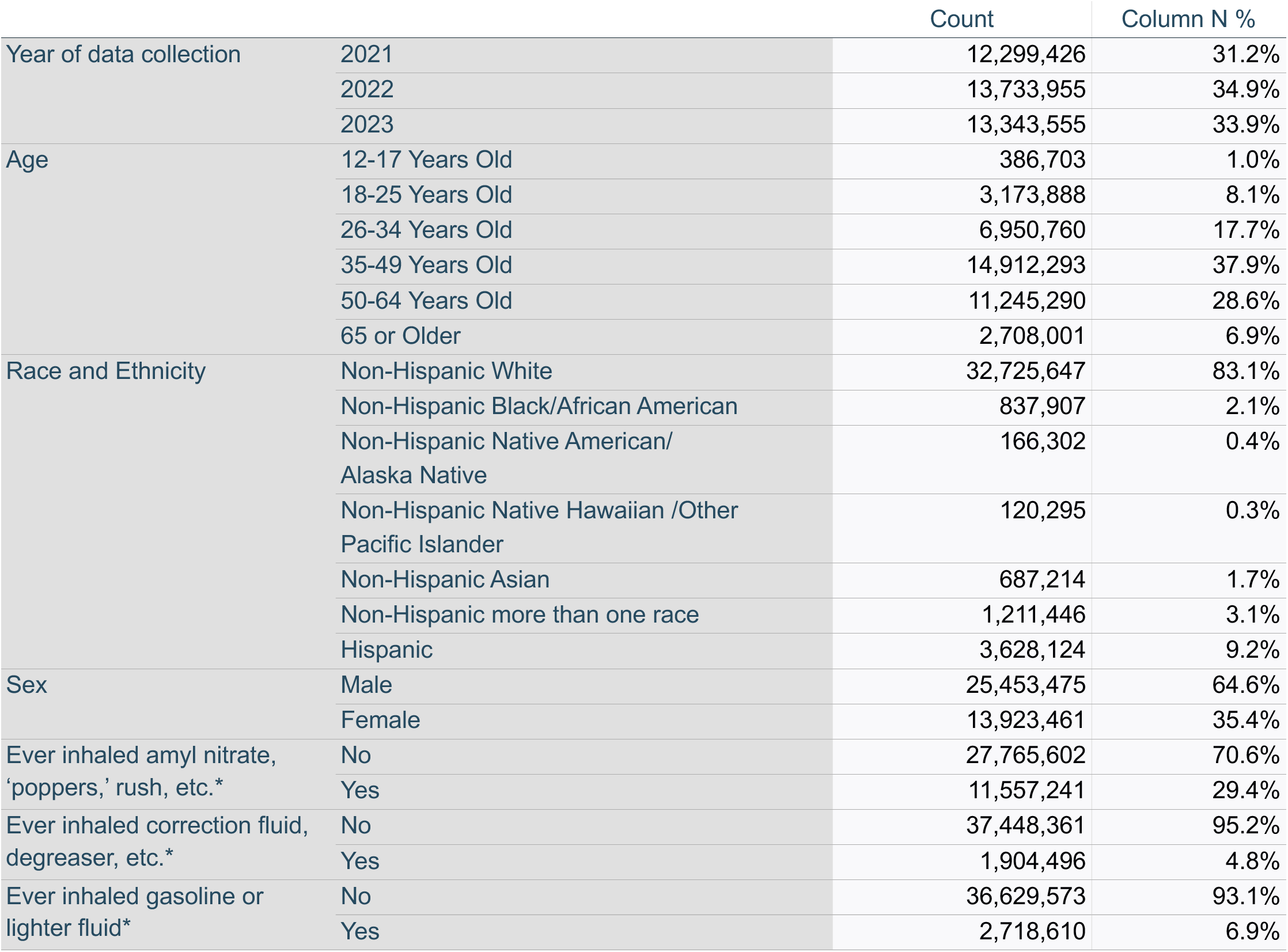
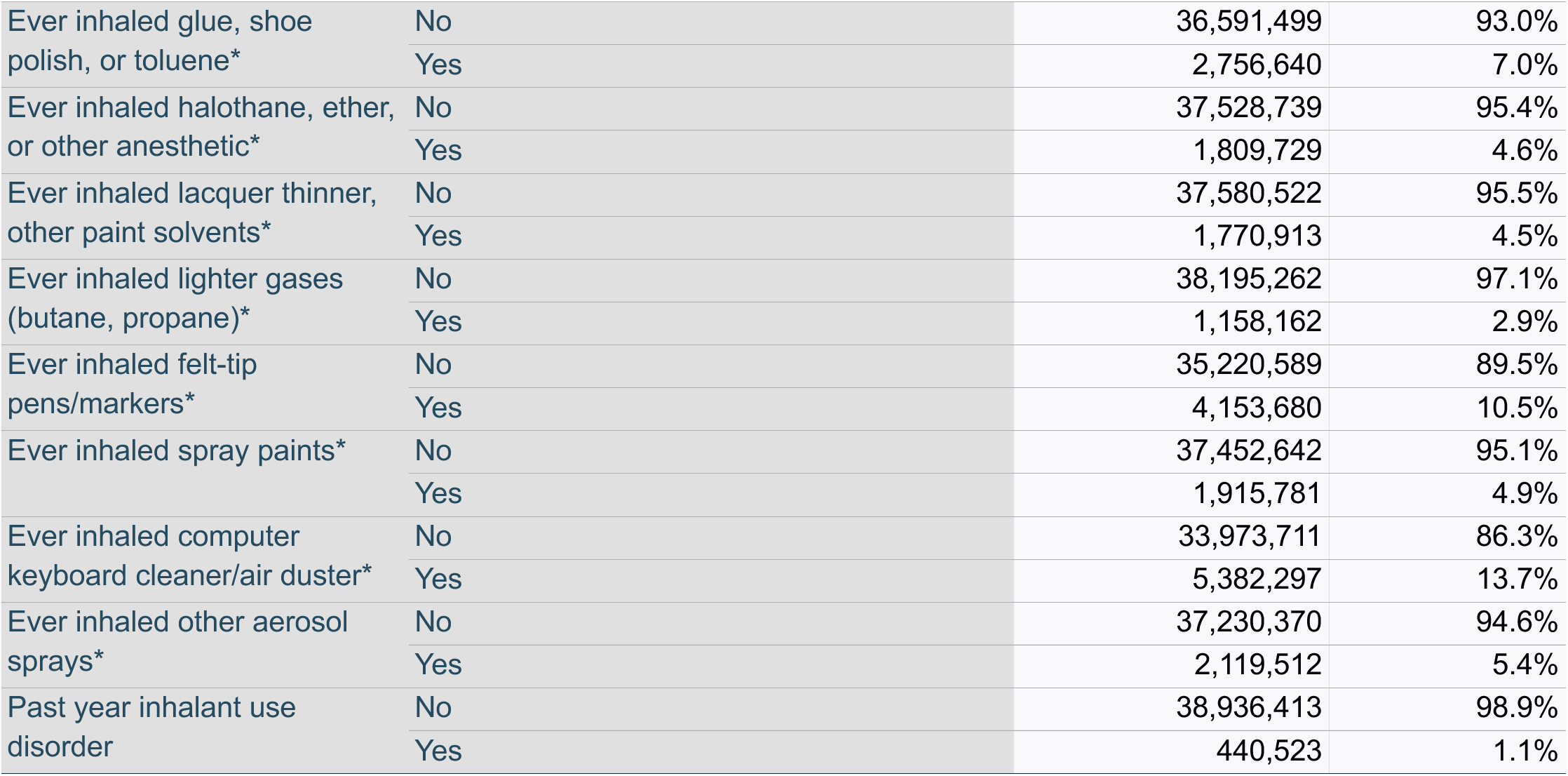
Aggregated Sample of Persons Reporting ever Using Nitrous Oxide ‘Whippits’ to get High using National Weighted Survey Data from 2021 to 2023; N = 39,374,736.

|  |  | Count | Column N % |
| --- | --- | --- | --- |
| Year of data collection | 2021 | 12,299,426 | 31.2% |
|  | 2022 | 13,733,955 | 34.9% |
|  | 2023 | 13,343,555 | 33.9% |
| Age | 12-17 Years Old | 386,703 | 1.0% |
|  | 18-25 Years Old | 3,173,888 | 8.1% |
|  | 26-34 Years Old | 6,950,760 | 17.7% |
|  | 35-49 Years Old | 14,912,293 | 37.9% |
|  | 50-64 Years Old | 11,245,290 | 28.6% |
|  | 65 or Older | 2,708,001 | 6.9% |
| Race and Ethnicity | Non-Hispanic White | 32,725,647 | 83.1% |
|  | Non-Hispanic Black/African American | 837,907 | 2.1% |
|  | Non-Hispanic Native American/<br>Alaska Native | 166,302 | 0.4% |
|  | Non-Hispanic Native Hawaiian /Other<br>Pacific Islander | 120,295 | 0.3% |
|  | Non-Hispanic Asian | 687,214 | 1.7% |
|  | Non-Hispanic more than one race | 1,211,446 | 3.1% |
|  | Hispanic | 3,628,124 | 9.2% |
| Sex | Male | 25,453,475 | 64.6% |
|  | Female | 13,923,461 | 35.4% |
| Ever inhaled amyl nitrate,<br>'poppers,' rush, etc.* | No | 27,765,602 | 70.6% |
|  | Yes | 11,557,241 | 29.4% |
| Ever inhaled correction fluid,<br>degreaser, etc.* | No | 37,448,361 | 95.2% |
|  | Yes | 1,904,496 | 4.8% |
| Ever inhaled gasoline or<br>lighter fluid* | No | 36,629,573 | 93.1% |
|  | Yes | 2,718,610 | 6.9% |
| Ever inhaled glue, shoe polish, or toluene* | No | 36,591,499 | 93.0% |
|  | Yes | 2,756,640 | 7.0% |
| Ever inhaled halothane, ether, or other anesthetic* | No | 37,528,739 | 95.4% |
|  | Yes | 1,809,729 | 4.6% |
| Ever inhaled lacquer thinner, other paint solvents* | No | 37,580,522 | 95.5% |
|  | Yes | 1,770,913 | 4.5% |
| Ever inhaled lighter gases (butane, propane)* | No | 38,195,262 | 97.1% |
|  | Yes | 1,158,162 | 2.9% |
| Ever inhaled felt-tip pens/markers* | No | 35,220,589 | 89.5% |
|  | Yes | 4,153,680 | 10.5% |
| Ever inhaled spray paints* | No | 37,452,642 | 95.1% |
|  | Yes | 1,915,781 | 4.9% |
| Ever inhaled computer keyboard cleaner/air duster* | No | 33,973,711 | 86.3% |
|  | Yes | 5,382,297 | 13.7% |
| Ever inhaled other aerosol sprays* | No | 37,230,370 | 94.6% |
|  | Yes | 2,119,512 | 5.4% |
| Past year inhalant use disorder | No | 38,936,413 | 98.9% |
|  | Yes | 440,523 | 1.1% |

Approximately 1.1% of the sample met the criteria for a past year inhalant use disorder (n = 440,523). Of the n = 440,523 individuals having an inhalant use disorder 337,612 had a mild inhalant use disorder, 51,451 had a moderate inhalant use disorder, and 51,460 had a severe inhalant use disorder. Table 2 presents annual counts and percentages for each year of data collection, 2021 to 2023. As shown in Table 2, Non-Hispanic White individuals and males constituted most of the sample in each year of data collection.

**Table 2.**
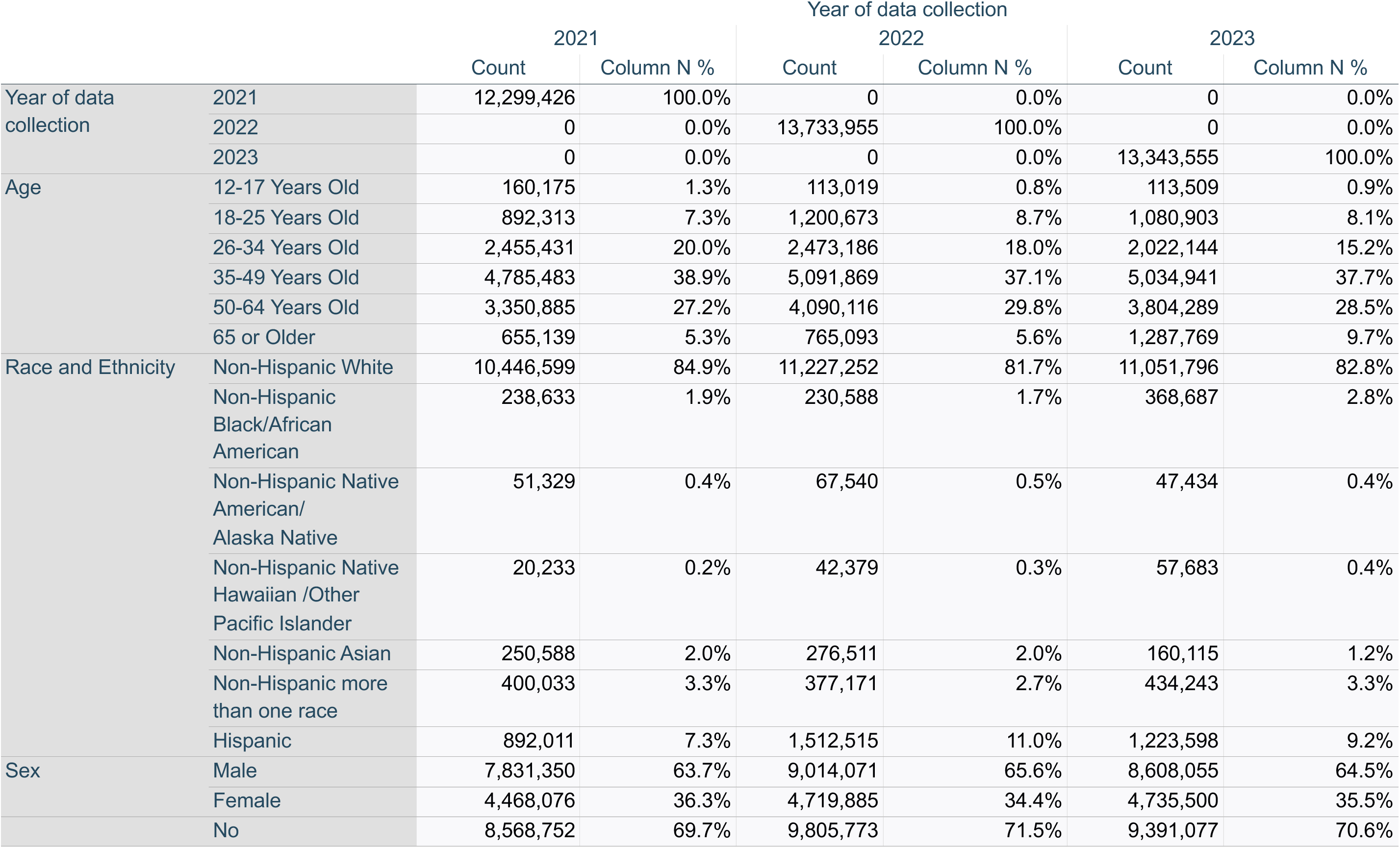
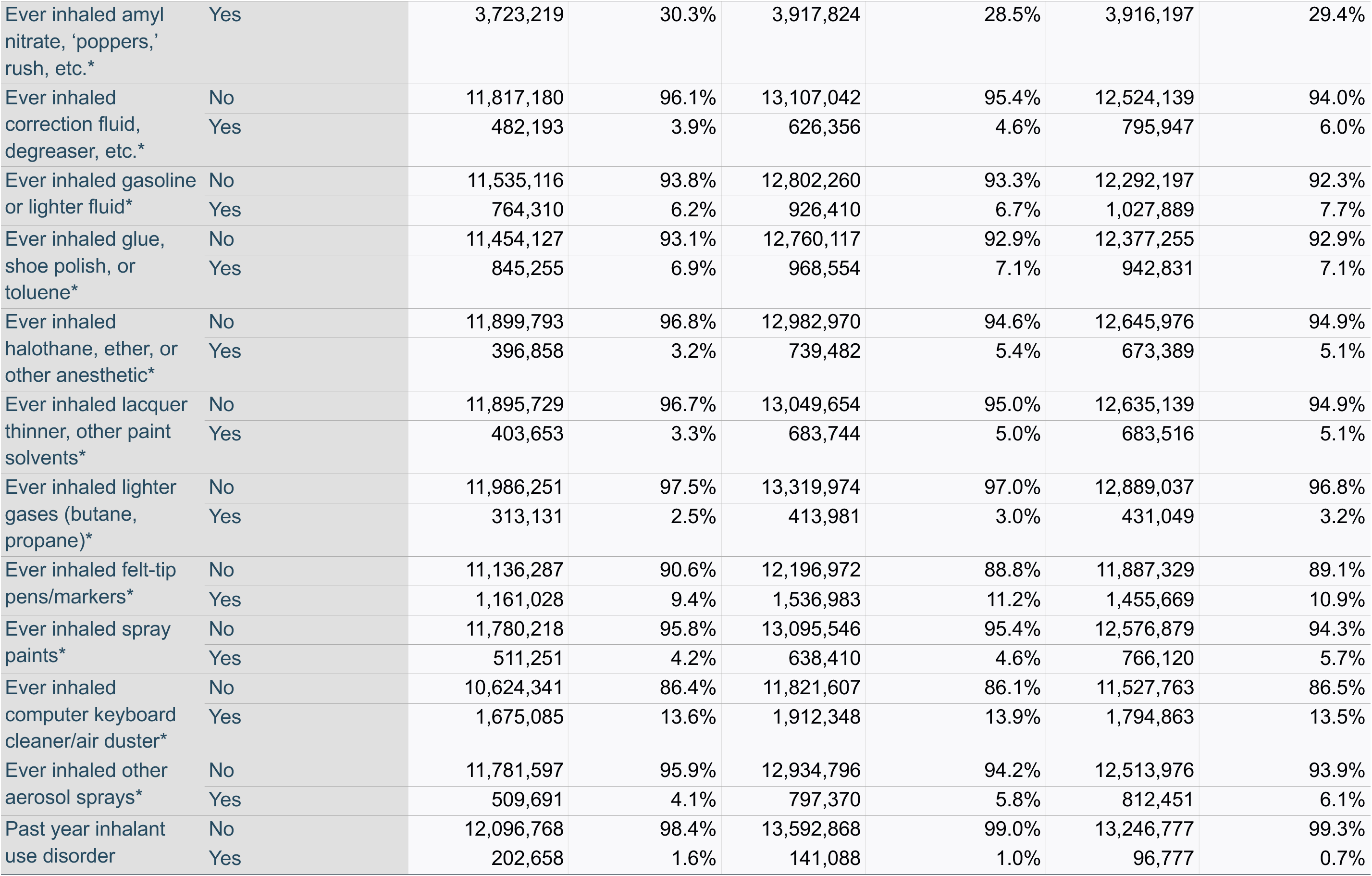
Annual Sample of Persons Reporting ever Using Nitrous Oxide ‘Whippits’ to get High using National Weighted Survey Data from 2021 to 2023; N = 39,374,736.

|  |  | Year of data collection |  |  |  |  |  |
| --- | --- | --- | --- | --- | --- | --- | --- |
|  |  | 2021 |  | 2022 |  | 2023 |  |
|  |  | Count | Column N % | Count | Column N % | Count | Column N % |
| Year of data collection | 2021 | 12,299,426 | 100.0% | 0 | 0.0% | 0 | 0.0% |
|  | 2022 | 0 | 0.0% | 13,733,955 | 100.0% | 0 | 0.0% |
|  | 2023 | 0 | 0.0% | 0 | 0.0% | 13,343,555 | 100.0% |
| Age | 12-17 Years Old | 160,175 | 1.3% | 113,019 | 0.8% | 113,509 | 0.9% |
|  | 18-25 Years Old | 892,313 | 7.3% | 1,200,673 | 8.7% | 1,080,903 | 8.1% |
|  | 26-34 Years Old | 2,455,431 | 20.0% | 2,473,186 | 18.0% | 2,022,144 | 15.2% |
|  | 35-49 Years Old | 4,785,483 | 38.9% | 5,091,869 | 37.1% | 5,034,941 | 37.7% |
|  | 50-64 Years Old | 3,350,885 | 27.2% | 4,090,116 | 29.8% | 3,804,289 | 28.5% |
|  | 65 or Older | 655,139 | 5.3% | 765,093 | 5.6% | 1,287,769 | 9.7% |
| Race and Ethnicity | Non-Hispanic White | 10,446,599 | 84.9% | 11,227,252 | 81.7% | 11,051,796 | 82.8% |
|  | Non-Hispanic Black/African American | 238,633 | 1.9% | 230,588 | 1.7% | 368,687 | 2.8% |
|  | Non-Hispanic Native American/ Alaska Native | 51,329 | 0.4% | 67,540 | 0.5% | 47,434 | 0.4% |
|  | Non-Hispanic Native Hawaiian /Other Pacific Islander | 20,233 | 0.2% | 42,379 | 0.3% | 57,683 | 0.4% |
|  | Non-Hispanic Asian | 250,588 | 2.0% | 276,511 | 2.0% | 160,115 | 1.2% |
|  | Non-Hispanic more than one race | 400,033 | 3.3% | 377,171 | 2.7% | 434,243 | 3.3% |
|  | Hispanic | 892,011 | 7.3% | 1,512,515 | 11.0% | 1,223,598 | 9.2% |
| Sex | Male | 7,831,350 | 63.7% | 9,014,071 | 65.6% | 8,608,055 | 64.5% |
|  | Female | 4,468,076 | 36.3% | 4,719,885 | 34.4% | 4,735,500 | 35.5% |
|  | No | 8,568,752 | 69.7% | 9,805,773 | 71.5% | 9,391,077 | 70.6% |
| Ever inhaled amyl nitrate, 'poppers,' rush, etc.* | Yes | 3,723,219 | 30.3% | 3,917,824 | 28.5% | 3,916,197 | 29.4% |
| Ever inhaled correction fluid, degreaser, etc.* | No | 11,817,180 | 96.1% | 13,107,042 | 95.4% | 12,524,139 | 94.0% |
|  | Yes | 482,193 | 3.9% | 626,356 | 4.6% | 795,947 | 6.0% |
| Ever inhaled gasoline or lighter fluid* | No | 11,535,116 | 93.8% | 12,802,260 | 93.3% | 12,292,197 | 92.3% |
|  | Yes | 764,310 | 6.2% | 926,410 | 6.7% | 1,027,889 | 7.7% |
| Ever inhaled glue, shoe polish, or toluene* | No | 11,454,127 | 93.1% | 12,760,117 | 92.9% | 12,377,255 | 92.9% |
|  | Yes | 845,255 | 6.9% | 968,554 | 7.1% | 942,831 | 7.1% |
| Ever inhaled halothane, ether, or other anesthetic* | No | 11,899,793 | 96.8% | 12,982,970 | 94.6% | 12,645,976 | 94.9% |
|  | Yes | 396,858 | 3.2% | 739,482 | 5.4% | 673,389 | 5.1% |
| Ever inhaled lacquer thinner, other paint solvents* | No | 11,895,729 | 96.7% | 13,049,654 | 95.0% | 12,635,139 | 94.9% |
|  | Yes | 403,653 | 3.3% | 683,744 | 5.0% | 683,516 | 5.1% |
| Ever inhaled lighter gases (butane, propane)* | No | 11,986,251 | 97.5% | 13,319,974 | 97.0% | 12,889,037 | 96.8% |
|  | Yes | 313,131 | 2.5% | 413,981 | 3.0% | 431,049 | 3.2% |
| Ever inhaled felt-tip pens/markers* | No | 11,136,287 | 90.6% | 12,196,972 | 88.8% | 11,887,329 | 89.1% |
|  | Yes | 1,161,028 | 9.4% | 1,536,983 | 11.2% | 1,455,669 | 10.9% |
| Ever inhaled spray paints* | No | 11,780,218 | 95.8% | 13,095,546 | 95.4% | 12,576,879 | 94.3% |
|  | Yes | 511,251 | 4.2% | 638,410 | 4.6% | 766,120 | 5.7% |
| Ever inhaled computer keyboard cleaner/air duster* | No | 10,624,341 | 86.4% | 11,821,607 | 86.1% | 11,527,763 | 86.5% |
|  | Yes | 1,675,085 | 13.6% | 1,912,348 | 13.9% | 1,794,863 | 13.5% |
| Ever inhaled other aerosol sprays* | No | 11,781,597 | 95.9% | 12,934,796 | 94.2% | 12,513,976 | 93.9% |
|  | Yes | 509,691 | 4.1% | 797,370 | 5.8% | 812,451 | 6.1% |
| Past year inhalant use disorder | No | 12,096,768 | 98.4% | 13,592,868 | 99.0% | 13,246,777 | 99.3% |
|  | Yes | 202,658 | 1.6% | 141,088 | 1.0% | 96,777 | 0.7% |

A sub-sample of cases was selected based on the following criteria: [a] lifetime use of nitrous oxide was reported, [b] past 12-month inhalant use was reported, and [c] the respondent never used other inhalants, which resulted in 1,271,122 cases. Among these cases that only used nitrous oxide and no other inhalants in the past 12 months, n = 130,339 (10.3% of this sub-sample) met the criteria for an inhalant use disorder from 2021 to 2023. Annually, 12.3% in 2021, 11.6% in 2022, and 6.5% in 2023 of this sub-sample met the criteria for an inhalant use disorder diagnosis attributable to nitrous oxide only since no other inhalant use was reported by this sub-sample.

### Nitrous Oxide Poisoning Data: 2021 to 2023

Annual count data of reported nitrous oxide poisonings may be found in Table 3. Across the entire analytic period of 2021 to 2023 there were 937 total poisoning cases in which nitrous oxide was involved, 712 poisoning cases in which nitrous oxide was the only substance involved in the exposure, and 507 of the cases were intentional exposures.

**Table 3.** Annual Nitrous Oxide Poison Data from 2021 to 2023.

| Year | Number of Case Mentions | Number of Single Exposures | Intentional Exposure |
| --- | --- | --- | --- |
| 2021 | 303 | 225 | 163 |
| 2022 | 283 | 211 | 145 |
| 2023 | 351 | 276 | 199 |

Data extracted from the Annual Report of the National Poison Data System® (NPDS) from America's Poison Centers® 2021 to 2023 data

## DISCUSSION

The recreational overuse of nitrous oxide is a public health issue.^24^ Although nitrous oxide has commercial uses in the food and fuel industries, some people misuse it for its psychoactive effects.^12, 13^ This study is the first to present national data on both lifetime use of nitrous oxide from a health survey and the total number of poisoning cases involving nitrous oxide from 2021 to 2023 in the U.S. Ultimately, about one in twenty Americans has used nitrous oxide recreationally.

Males constituted the majority of the sample, which matches other research on nitrous oxide;^9^ however, the psychoactive effects of nitrous oxide seem to be similar across sexes.^2, 32^ Based on existing studies, adolescents and young adults are identified as the most likely to misuse nitrous oxide.^4, 9, 24^ Since the largest age groups in this study were between 35 to 49, followed by 50 to 64, this may suggest that people who have used nitrous oxide recreationally tend to be older.

Time will tell if these trends persist with current youth, especially since vaping has now become a public health concern among younger populations.^33-35^ While this does not mean that using one substance prevents the use of another, as polysubstance use is common, different generations generally favor certain psychoactive substances more than others.

This study examined the presence of past-year inhalant use disorder, which may also include meeting diagnostic criteria due to other inhalants used, such as lighter gases or spray paints. Of this sample of individuals who ever reported using nitrous oxide, over nine out of ten have not used an inhalant in the past 12 months. However, in the past year, inhalant use disorder was identified in 1.1% of the overall sample and in 1.6%, 1.0%, and 0.7% of the sample annually from 2021 to 2023. Among individuals who only used nitrous oxide in their lifetime to the exclusion of all other inhalants, and who reported using inhalants in the past year, approximately 10% met the criteria for inhalant use disorder due to their nitrous oxide use. This study builds on literature suggesting that individuals could meet the criteria for a substance use disorder due to their nitrous oxide use.^2, 9^ In the NSDUH, because nitrous oxide is included in the inhalant category, the questionnaire does not assess withdrawal. Withdrawal is not a commonly recognized symptom related to a substance use disorder with nitrous oxide;^3, 22^ however, other symptoms such as craving and tolerance have been identified.^2, 3^ Future national studies are needed to examine the prevalence of individuals meeting the criteria for a substance use disorder solely due to their nitrous oxide use, even if they co-use other inhalants. Background literature suggests that while nitrous oxide use occurs in both rural and urban settings, it may be more common in rural areas.^4^ Future research should investigate differences in the prevalence of inhalant use disorders between urban and rural contexts.

Nitrous oxide related poisonings were also examined in this current study. While the intentional cases included in this study also incorporate those in which self-harm was the purpose, deaths related to problematic nitrous oxide use in which self-harm was not intended also occur.^4^ This is an important consideration as other studies have highlighted several adverse events and hospitalizations related to problematic nitrous oxide use.^9, 17, 24^ This study identified 937 total poisoning cases in which nitrous oxide was involved, with 507 of the cases being intentional exposures. A study examining Centers for Disease Control and Prevention death data identified 1,240 deaths from 2010 to 2023 with an increase of identified cases during this analytic period.^23^ Further, the use of nitrous oxide was a contributing factor in 156 deaths identified in 2023.^23^ Taken together, this study continues to build on the literature of potential harms related to recreational nitrous oxide use.^36^ Due to the lack of screening tools to identify nitrous oxide use and healthcare providers probably not considering it during screening, nitrous oxide misuse may not always be identified in healthcare settings^4, 12^ unless prompted by the care recipient during self-disclosure of use.^13^ Problematic and long-term use of nitrous oxide can be associated with several harms, such as both acute and chronic neurological consequences, including paresthesia, neuropathy, and myelopathy.^3, 4, 10, 13, 16, 17^ Given the potential of nitrous oxide to reduce vitamin B12, B12 supplementation is recommended as a treatment regimen.^13^

### Limitations

Although nitrous oxide use can be diagnostically classified as “other substance use disorder”,^3, 9^ and this current study examines the prevalence of inhalant use disorder, nitrous oxide is listed as an inhalant in the NSDUH and in a large real-world treatment dataset based in the United States called the Treatment Episode Data Set Admissions.^20, 21^ This study also examined inhalant use disorder overall, which also includes persons who may meet diagnostic criteria based on using other inhalants instead of nitrous oxide. However, the sub-sample of individuals who used only nitrous oxide and no other inhalants was examined to provide rough estimates of inhalant use disorder attributable to nitrous oxide in this sub-sample. Considering that many individuals use multiple substances, this is not generalizable to persons who have used nitrous oxide and at least one other inhalant in their lifetime. Biases related to self-reports are also a limitation of the NSDUH. As a household survey, the NSDUH does not provide representative data on individuals who may be unhoused. The Annual Report of the National Poison Data System® (NPDS) from America’s Poison Centers® data does not capture all nitrous oxide poisonings, but only captures reported poisonings. Therefore, the number of total nitrous oxide poisonings could be greater.

## CONSLUSION

Problematic nitrous oxide use can result in harm. This descriptive study, conducted in the U.S. from 2021 to 2023, examined the prevalence of ever using nitrous oxide, assessed the proportion of individuals who met the past-year criteria for an inhalant use disorder, and reported the annual number of nitrous oxide poisonings. From 2021 to 2023, one in twenty persons in the U.S. reported ever using nitrous oxide recreationally. Past year inhalant use disorder among persons who ever used nitrous oxide recreationally was 1.1%. However, the inhalant use disorder diagnosis could be due to the use of another inhalant such as poppers. Among individuals who only used nitrous oxide in their lifetime to the exclusion of all other inhalants, and who reported using inhalants in the past year, approximately 10% met the criteria for inhalant use disorder due to their nitrous oxide use. From 2021 to 2023 there were 937 total poisoning cases in which nitrous oxide was involved with 712 cases involving nitrous oxide as the only reported substance. More population-level studies are needed to examine the national harms of problematic nitrous oxide use.

## Data Availability

De-identified data were used from the U.S.-based National Survey on Drug Use and Health and Annual Report of the National Poison Data System® (NPDS) from America’s Poison Centers®. These datasets may be found using the following links: [1] https://www.samhsa.gov/data/data-we-collect/nsduh-national-survey-drug-use-and-health/national-releases [2] https://doi.org/10.1080/15563650.2022.2132768 [3] https://doi.org/10.1080/15563650.2023.2268981 [4] https://doi.org/10.1080/15563650.2024.2412423.

## Ethical considerations

Study procedures were considered not human subjects research based on the University of North Carolina at Chapel Hill Institutional Review Board.

## Informed Consent

NOT APPLICABLE. Consent is not applicable as this study used publicly available data.

